# Evaluation of an artificial intelligence model for detection of pneumothorax and tension pneumothorax on chest radiograph

**DOI:** 10.1101/2022.07.07.22277305

**Authors:** James M Hillis, Bernardo C Bizzo, Sarah Mercaldo, John K Chin, Isabella Newbury-Chaet, Subba R Digumarthy, Matthew D Gilman, Victorine V Muse, Georgie Bottrell, Jarrel CY Seah, Catherine M Jones, Mannudeep K Kalra, Keith J Dreyer

## Abstract

**Importance:** Early detection of pneumothorax, most often on chest radiograph (CXR), can help determine need for emergent clinical intervention. The ability to accurately detect and rapidly triage pneumothorax with an artificial intelligence (AI) model could assist with earlier identification and improve care.

**Objective:** This study aimed to compare the accuracy of an AI model (Annalise Enterprise) to consensus thoracic radiologist interpretations in detecting (1) pneumothorax (incorporating both non-tension and tension pneumothorax) and (2) tension pneumothorax.

**Design:** A retrospective standalone performance assessment was conducted on a dataset of 1,000 CXR cases.

**Setting:** The cases were obtained from four hospitals in the United States.

**Participants:** The cases were obtained from patients aged 18 years or older. They were selected using two strategies from all CXRs performed at the hospitals including inpatients and outpatients. The first strategy identified consecutive pneumothorax cases through a manual review of radiology reports and the second strategy identified consecutive tension pneumothorax cases using natural language processing. For both strategies, negative cases were selected by taking the next negative case acquired from the same x-ray machine. The final dataset was an amalgamation of these processes.

**Methods:** Each case was interpreted independently by up to three radiologists to establish consensus ground truth interpretations. Each case was then interpreted by the AI model for the presence of pneumothorax and tension pneumothorax.

**Main Outcome:** The primary endpoints were the areas under the receiver operating characteristic curves (AUCs) for the detection of pneumothorax and tension pneumothorax. The secondary endpoints were the sensitivities and specificities for the detection of pneumothorax and tension pneumothorax at predefined operating points.

**Results:** Model inference was successfully performed in 307 non-tension pneumothorax, 128 tension pneumothorax and 550 negative cases. The AI model detected pneumothorax with AUC of 0.979 (94.3% sensitivity, 92.0% specificity) and tension pneumothorax with AUC of 0.987 (94.5% sensitivity, 95.3% specificity).

**Conclusions and Relevance:** The assessed AI model accurately detected pneumothorax and tension pneumothorax on this CXR dataset. Its use in the clinical workflow could lead to earlier identification and improved care for patients with pneumothorax.

**Key Points:** *Question:* Does a commercial artificial intelligence model accurately detect simple and tension pneumothorax on chest x-ray?

*Findings:* This retrospective study used 1,000 chest x-rays from four hospitals in the United States to compare artificial intelligence model outputs to consensus thoracic radiologist interpretations. The model detected pneumothorax (incorporating both simple and tension pneumothorax) with area under the curve (AUC) of 0.979 and tension pneumothorax with AUC of 0.987. The sensitivity and specificity were 94.3% and 92.0% respectively for pneumothorax, and 94.5% and 95.3% for tension pneumothorax.

*Meaning:* This artificial intelligence model could assist radiologists through its accurate detection of pneumothorax.

## Introduction

The diagnosis of a pneumothorax, especially tension pneumothorax, on chest radiograph (CXR) can lead to critical interventions for patient care.^1^ The automated detection of pneumothorax and tension pneumothorax through artificial intelligence (AI) has been hypothesized to improve patient care in multiple ways.^2,3^ Firstly, it can assist in triaging CXRs for sooner interpretation by a radiologist based on the suspected presence of a pneumothorax. Secondly, it can provide a second “set of eyes” to support identification of a pneumothorax.

Several AI models have been developed to assess for the presence of pneumothorax.^4-9^ Many models have also received US Food and Drug Administration (FDA) clearance for use in pneumothorax identification.^10-17^ These cleared models are all computer assisted triage devices, which are intended to aid in prioritization and triage of time sensitive findings.^18^

This retrospective study aimed to assess a commercial AI model that detects both pneumothorax and tension pneumothorax. It examined the model performance across a range of technical and demographic subgroups to assess the generalizability of the model. It also examined the model performance in the presence or absence of ancillary findings to determine how they impact model accuracy.

## Methods

### Study design

This retrospective standalone model performance study was conducted using radiology cases from four hospitals within the Mass General Brigham (MGB) network. It was approved by the MGB Institutional Review Board with waiver of informed consent. It was conducted in accordance with relevant guidelines and regulations including the Health Insurance Portability and Accountability Act (HIPAA).

### Case selection

The cohort was selected based on two strategies that utilized the radiology reports within the MGB radiology archive: consecutive pneumothorax cases through manual review and consecutive tension pneumothorax cases through a natural language processing search engine. Each strategy involved taking the next negative case acquired on the same x-ray machine after each positive case to avoid temporal and technical bias. The cohort considered all CXRs performed at a hospital including inpatient and outpatient. The CXRs were obtained from patients at least 18 years of age.

For the consecutive pneumothorax cases, the consecutive cases were identified in a prospective order between June 1 2019 and May 31 2021. There were 85 report-positive cases and 85 report-negative cases from each of the four hospitals. A radiologist reviewed consecutive CXR reports at each of the hospitals until these numbers were achieved.

For the consecutive tension pneumothorax cases, the report-positive cases were identified between June 1 2015 and May 31 2021 using a natural language processing search for “tension pneumothorax” amongst CXR reports from the same four hospitals utilizing a commercial radiology report search engine (Nuance mPower Clinical Analytics). A radiologist then confirmed these cases were positive through a CXR report review. There were 160 report-positive cases selected across all four hospitals in a consecutive, retrospective manner from the most recent. An equal number of report-negative cases were then selected by taking the next report-negative case after each report-positive case on each x-ray machine.

All cases were deidentified and underwent an image quality review by an American Board of Radiology (ABR)-certified radiologist. The image quality review identified the patient positioning (erect or supine) and projections present (antero-posterior (AP), postero-anterior (PA) and/or lateral). Cases were excluded if they did not include a CXR or did not include a frontal projection (AP or PA). The review was performed using the FDA-cleared eUnity image visualization software (Version 6 or higher) and an internal web-based annotation system.

### Ground truth interpretations

Ground truth interpretations were performed by three ABR-certified radiologists with fellowship training in thoracic radiology. They provided their interpretations independently, without access to the original radiology reports and in different worklist orders. They used the same image visualization software and annotation system as was used in the image quality review.

The radiologists answered one or three multiple choice questions about pneumothorax: if a pneumothorax was present or absent, if the size of pneumothorax was <2cm or ≥2cm, and if features of tension pneumothorax were present or absent (the latter two questions only applied if a pneumothorax was present). They also indicated the presence of ten ancillary findings: pleural effusion (including hemothorax), rib fracture, pneumomediastinum, pneumoperitoneum, subcutaneous emphysema, focal or diffuse pulmonary abnormalities (including nodules, masses, emphysema, airspace or interstitial processes), skin fold, intercostal drain, evidence of thoracic surgery (including thoracotomy wires or sutures), and other lines / tubes / devices (including pacemaker, endotracheal tube, central line).

For determining consensus for the three pneumothorax findings, a “2+1” strategy was used: the first two radiologists interpreted every case and a third radiologist then interpreted cases with discrepant interpretations. Any persistent discrepancies (which could occur for the size and features of tension pneumothorax questions after the three interpretations) were resolved at a meeting of all three radiologists. The ancillary findings were considered present if any radiologist interpreted them as being present.

### Model inference

The evaluated AI model was version 2.0.0 of the Annalise Enterprise CXR Triage Pneumothorax device. This device is a variant of the Annalise Enterprise (CXR module) device, which is commercially available in some non-US markets. The Annalise Enterprise (CXR module) device is trained to identify over 100 different radiological findings and is a deep-convolutional neural network trained on over 750,000 CXRs, which were each labelled by three radiologists.^19^ The model was installed at MGB for use in this study and received only the Digital Imaging and Communications in Medicine (DICOM)-formatted CXR cases.

### Analysis

The statistical analysis was performed in R (version 4.0.2) on the full analysis set. The predefined primary endpoints were the areas under the receiver operating characteristic curves (AUCs) for the detection of pneumothorax (incorporating non-tension and tension pneumothorax) and tension pneumothorax; they were calculated using the consensus annotations and the classification scores from the AI model. The predefined secondary endpoints were the sensitivities and specificities for the detection of pneumothorax and tension pneumothorax; they were calculated using the same outputs as the AUCs and the operating points of the model.

These analyses were repeated for additional subgroups as detailed in the results. The sex, age and manufacturers were derived from clinical databases or DICOM fields for each radiology case. Any missing data were treated as “Unknown” and no data were imputed. The manufacturers in the DICOM field may have represented manufacturers other than the x-ray machine (e.g., the cassette). The patient positioning, projections and ancillary findings were obtained as part of the case selection or ground truth interpretations as described above. All confidence intervals (CIs) were calculated using bootstrapped intervals with 2,000 resamples. The predefined passing criterion for the primary endpoints was AUC >0.95 based on recognized FDA performance benchmarks.^18^ While not predefined passing criteria, this manuscript refers to additional benchmarks of sensitivity >80% and specificity >80%. The sample sizes for each of pneumothorax and tension pneumothorax were calculated using prior model results and to ensure the lower bound of the 95% CI for AUC was >0.95.

## Results

### Participants

An initial cohort of 1,000 CXR cases were selected for this project (Supplementary Figure 1). Three cases were excluded as they did not contain a CXR during an image quality review. Twelve cases were unsuccessful at model inference. The remaining 985 cases were used for analysis. They included 435 (44.2%) positive pneumothorax cases and 550 (55.8%) negative pneumothorax cases. There were 128 cases that were positive for tension pneumothorax (29.4% of positive pneumothorax cases; 13.0% of total cases). The demographic and technical breakdowns are provided in Table 1.

**Table 1:** Demographic and technical breakdown of CXR cases

|  | Positive for pneumothorax |  |  | Negative for pneumothorax |
| --- | --- | --- | --- | --- |
|  | Positive for tension pneumothorax | Negative for tension pneumothorax | Total | Total |
| **Total** | 128 | 307 | 435 | 550 |
| **Sex** |  |  |  |  |
| Female | 56 (43.8%) | 125 (40.7%) | 181 (41.6%) | 255 (46.4%) |
| Male | 72 (56.2%) | 182 (59.3%) | 254 (58.4%) | 295 (53.6%) |
| **Age** |  |  |  |  |
| ≤65 | 80 (62.5%) | 154 (50.2%) | 234 (53.8%) | 289 (52.5%) |
| >65 | 48 (37.5%) | 153 (49.8%) | 201 (46.2%) | 261 (47.5%) |
| Mean (years) | 57.5 ± 18.9 | 60.2 ± 19.4 | 59.4 ± 19.3 | 61.9 ± 18.7 |
| **Manufacturer** |  |  |  |  |
| Agfa | 70 (54.7%) | 91 (29.6%) | 161 (37.0%) | 194 (35.3%) |
| Carestream | 4 (3.1%) | 37 (12.1%) | 41 (9.4%) | 47 (8.5%) |
| Fujifilm | 7 (5.5%) | 24 (7.8%) | 31 (7.1%) | 74 (13.5%) |
| GE Healthcare | 0 (0.0%) | 3 (1.0%) | 3 (0.7%) | 7 (1.3%) |
| Kodak | 0 (0.0%) | 7 (2.3%) | 7 (1.6%) | 14 (2.5%) |
| Konica Minolta | 24 (18.8%) | 38 (12.4%) | 62 (14.3%) | 78 (14.2%) |
| McKesson | 7 (5.5%) | 30 (9.8%) | 37 (8.5%) | 13 (2.4%) |
| Philips | 5 (3.9%) | 43 (14.0%) | 48 (11.0%) | 52 (9.5%) |
| Siemens | 5 (3.9%) | 15 (4.9%) | 20 (4.6%) | 32 (5.8%) |
| Varian | 6 (4.7%) | 17 (5.5%) | 23 (5.3%) | 26 (4.7%) |
| Unknown | 0 (0.0%) | 2 (0.7%) | 2 (0.5%) | 13 (2.4%) |
| **Patient Positioning** |  |  |  |  |
| Erect | 69 (53.9%) | 221 (72.0%) | 290 (66.7%) | 370 (67.3%) |
| Supine | 59 (46.1%) | 86 (28.0%) | 145 (33.3%) | 180 (32.7%) |
| **Projections (can include multiple)** |  |  |  |  |
| AP | 111 (86.7%) | 202 (65.8%) | 313 (72.0%) | 386 (70.2%) |
| PA | 17 (13.3%) | 105 (34.2%) | 122 (28.0%) | 164 (29.8%) |
| Lateral | 20 (15.6%) | 104 (33.9%) | 124 (28.5%) | 177 (32.2%) |
| **Pneumothorax Size** |  |  |  |  |
| <2cm | 0 (0.0%) | 164 (53.4%) | 164 (37.7%) | N/A |
| ≥2cm | 128 (100.0%) | 143 (46.6%) | 271 (62.3%) | N/A |

### Pneumothorax detection

The AI model identified pneumothorax (incorporating non-tension and tension pneumothorax) with AUC 0.979 (95% CI 0.970-0.987; Figure 1A), sensitivity 94.3% (95% CI 92.0-96.3%) and specificity 92.0% (95% CI 89.6-94.2%). When considering the subgroup analyses for sex, age, manufacturer, patient positioning and projections, all subgroups achieved at least AUC 0.95, sensitivity 80% and specificity 80% except the Kodak and Unknown manufacturer subgroups (Table 2). These two subgroups had small cohort sizes and underperformed due to false negative cases: 2 false negative cases out of 7 total positive cases for the Kodak manufacturer subgroup and 1 false negative case out of 2 total positive cases for the Unknown manufacturer subgroup. The model achieved higher sensitivity for detecting pneumothorax amongst tension (compared with non-tension) pneumothorax and ≥2cm (compared with <2cm) sized pneumothorax. It detected a pneumothorax in all cases that had a tension pneumothorax.

**Figure 1:**
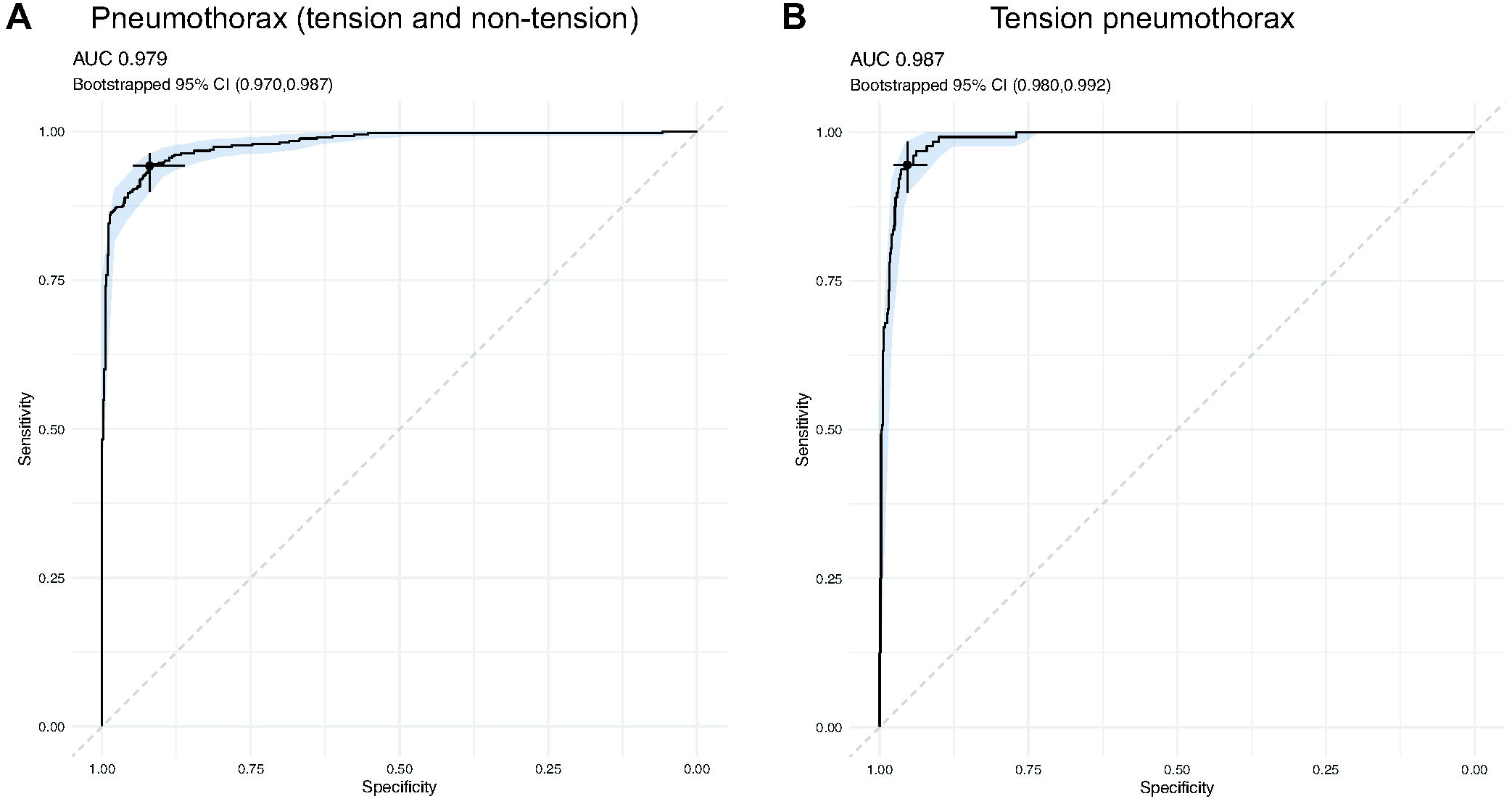
Receiver operating characteristic curve for pneumothorax and tension pneumothorax detection: Graphs demonstrate the ability of the AI model to detect pneumothorax (A) and tension pneumothorax (B). The shaded region reflects the bootstrapped 95% confidence interval. The selected point on each graph reflects the model operating point. AUC: area under the curve, CI: confidence interval.

**Table 2:** Performance for detecting pneumothorax across demographic and technical subgroups

|  | Positive N | Negative N | AUC (95% CI) | Sensitivity (95% CI) | Specificity (95% CI) |
| --- | --- | --- | --- | --- | --- |
| **Overall** | 435 | 550 | 0.979 (0.970-0.987) | 94.3% (92.0-96.3%) | 92.0% (89.6-94.2%) |
| **Sex** |  |  |  |  |  |
| Female | 181 | 255 | 0.978 (0.963-0.989) | 92.8% (89.0-96.1%) | 92.2% (88.6-95.3%) |
| Male | 254 | 295 | 0.981 (0.971-0.989) | 95.3% (92.5-97.6%) | 91.9% (88.8-94.9%) |
| **Age** |  |  |  |  |  |
| ≤65 | 234 | 289 | 0.983 (0.973-0.990) | 95.7% (93.2-97.9%) | 90.7% (87.2-93.8%) |
| >65 | 201 | 261 | 0.976 (0.961-0.988) | 92.5% (88.6-96.0%) | 93.5% (90.4-96.2%) |
| **Manufacturer** |  |  |  |  |  |
| Agfa | 161 | 194 | 0.974 (0.958-0.985) | 94.4% (90.7-97.5%) | 85.6 (80.4-90.2%) |
| Carestream | 41 | 47 | 0.988 (0.960-1.000) | 95.1% (87.8-100.0%) | 91.5 (83.0-97.9%) |
| Fujifilm | 31 | 74 | 0.998 (0.991-1.000) | 96.8% (90.3-100.0%) | 95.9 (90.5-100.0%) |
| GE Healthcare | 3 | 7 | 1.000 (1.000-1.000) | 100.0% (100.0-100.0%) | 100.0 (100.0-100.0%) |
| Kodak | 7 | 14 | 0.898 (0.663-1.000) | 71.4% (42.9-100.0%) | 100.0 (100.0-100.0%) |
| Konica Minolta | 62 | 78 | 0.973 (0.943-0.993) | 96.8% (91.9-100.0%) | 89.7 (82.1-96.2%) |
| McKesson | 37 | 13 | 1.000 (1.000-1.000) | 97.3% (91.9-100.0%) | 100.0 (100.0-100.0%) |
| Philips | 48 | 52 | 0.988 (0.967-0.998) | 89.6% (79.2-97.9%) | 100.0 (100.0-100.0%) |
| Siemens | 20 | 32 | 1.000 (1.000-1.000) | 100.0% (100.0-100.0%) | 96.9 (90.6-100.0%) |
| Varian | 23 | 26 | 0.990 (0.955-1.000) | 91.3% (78.3-100.0%) | 100.0 (100.0-100.0%) |
| Unknown | 2 | 13 | 1.000 (1.000-1.000) | 50.0% (0.0-100.0%) | 100.0 (100.0-100.0%) |
| **Patient Positioning** |  |  |  |  |  |
| Erect | 290 | 370 | 0.982 (0.972-0.990) | 93.1% (90.0-95.9%) | 94.6% (92.2-96.8%) |
| Supine | 145 | 180 | 0.976 (0.960-0.988) | 96.6% (93.1-99.3%) | 86.7% (81.7-91.7%) |
| **Projections** |  |  |  |  |  |
| AP/PA without lateral | 311 | 373 | 0.977 (0.968-0.985) | 95.2% (92.6-97.4%) | 88.5% (85.3-91.4%) |
| AP/PA with lateral | 124 | 177 | 0.988 (0.971-0.997) | 91.9% (87.1-96.8%) | 99.4% (98.3-100.0%) |
| **Features of tension** |  |  |  |  |  |
| Absent | 307 | N/A | N/A | 91.9% (88.6-94.8%) | N/A |
| Present | 128 | N/A | N/A | 100.0% (100.0-100.0%) | N/A |
| **Pneumothorax Size** |  |  |  |  |  |
| <2cm | 164 | N/A | N/A | 86.6% (81.1-91.5%) | N/A |
| ≥2cm | 271 | N/A | N/A | 98.9% (97.4-100.0%) | N/A |

### Tension pneumothorax detection

The AI model identified tension pneumothorax with AUC 0.987 (95% CI 0.980-0.992; Figure 1B), sensitivity 94.5% (95% CI 90.6-97.7%) and specificity 95.3% (95% CI 93.9-96.6%). When considering the subgroup analyses for sex, age, manufacturer, patient positioning and projections, all subgroups achieved at least AUC 0.95, sensitivity 80% and specificity 80% (Table 3).

**Table 3:**
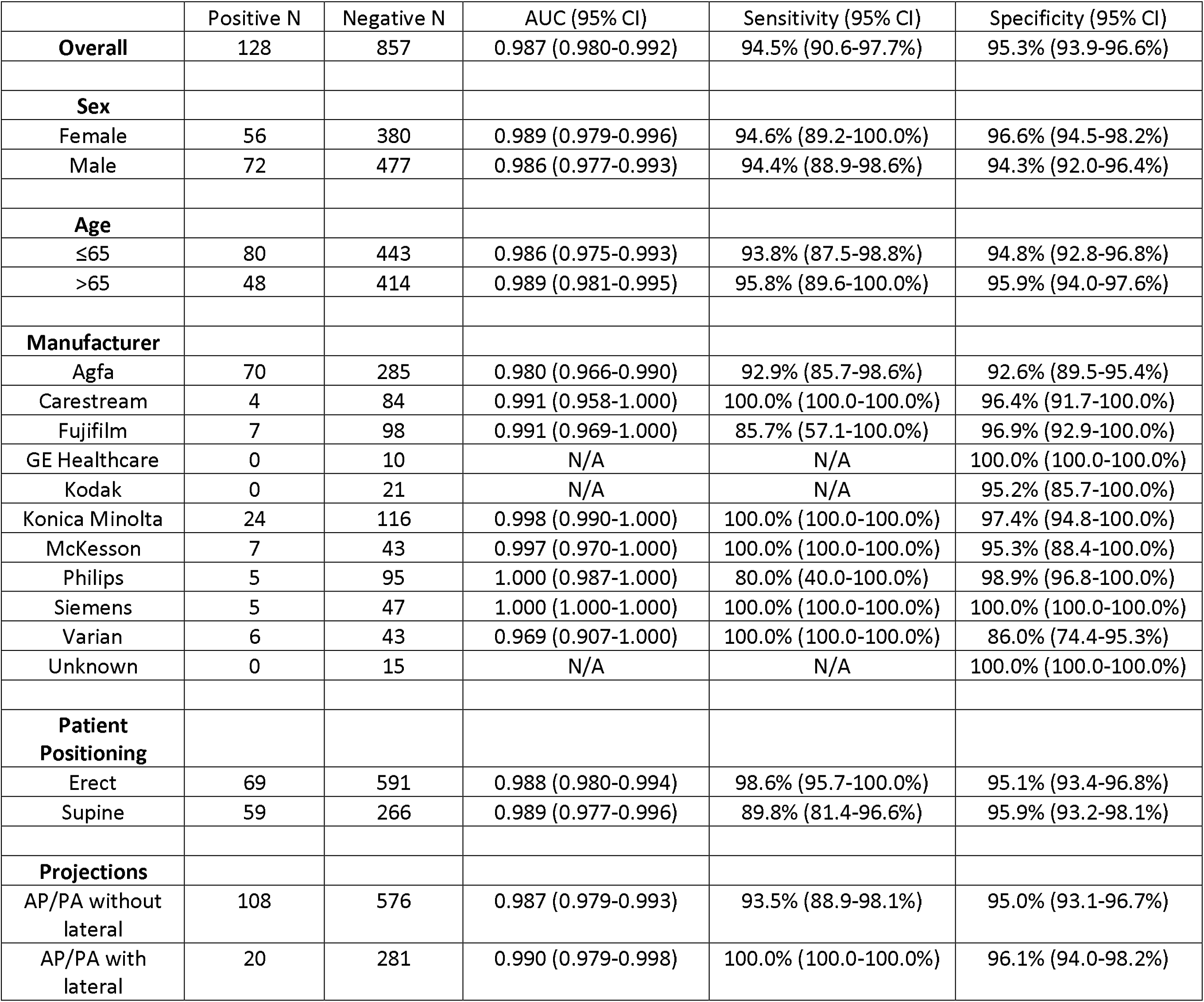
Performance for detecting tension pneumothorax across demographic and technical subgroups

|  | Positive N | Negative N | AUC (95% CI) | Sensitivity (95% CI) | Specificity (95% CI) |
| --- | --- | --- | --- | --- | --- |
| **Overall** | 128 | 857 | 0.987 (0.980-0.992) | 94.5% (90.6-97.7%) | 95.3% (93.9-96.6%) |
| **Sex** |  |  |  |  |  |
| Female | 56 | 380 | 0.989 (0.979-0.996) | 94.6% (89.2-100.0%) | 96.6% (94.5-98.2%) |
| Male | 72 | 477 | 0.986 (0.977-0.993) | 94.4% (88.9-98.6%) | 94.3% (92.0-96.4%) |
| **Age** |  |  |  |  |  |
| ≤65 | 80 | 443 | 0.986 (0.975-0.993) | 93.8% (87.5-98.8%) | 94.8% (92.8-96.8%) |
| >65 | 48 | 414 | 0.989 (0.981-0.995) | 95.8% (89.6-100.0%) | 95.9% (94.0-97.6%) |
| **Manufacturer** |  |  |  |  |  |
| Agfa | 70 | 285 | 0.980 (0.966-0.990) | 92.9% (85.7-98.6%) | 92.6% (89.5-95.4%) |
| Carestream | 4 | 84 | 0.991 (0.958-1.000) | 100.0% (100.0-100.0%) | 96.4% (91.7-100.0%) |
| Fujifilm | 7 | 98 | 0.991 (0.969-1.000) | 85.7% (57.1-100.0%) | 96.9% (92.9-100.0%) |
| GE Healthcare | 0 | 10 | N/A | N/A | 100.0% (100.0-100.0%) |
| Kodak | 0 | 21 | N/A | N/A | 95.2% (85.7-100.0%) |
| Konica Minolta | 24 | 116 | 0.998 (0.990-1.000) | 100.0% (100.0-100.0%) | 97.4% (94.8-100.0%) |
| McKesson | 7 | 43 | 0.997 (0.970-1.000) | 100.0% (100.0-100.0%) | 95.3% (88.4-100.0%) |
| Philips | 5 | 95 | 1.000 (0.987-1.000) | 80.0% (40.0-100.0%) | 98.9% (96.8-100.0%) |
| Siemens | 5 | 47 | 1.000 (1.000-1.000) | 100.0% (100.0-100.0%) | 100.0% (100.0-100.0%) |
| Varian | 6 | 43 | 0.969 (0.907-1.000) | 100.0% (100.0-100.0%) | 86.0% (74.4-95.3%) |
| Unknown | 0 | 15 | N/A | N/A | 100.0% (100.0-100.0%) |
| **Patient Positioning** |  |  |  |  |  |
| Erect | 69 | 591 | 0.988 (0.980-0.994) | 98.6% (95.7-100.0%) | 95.1% (93.4-96.8%) |
| Supine | 59 | 266 | 0.989 (0.977-0.996) | 89.8% (81.4-96.6%) | 95.9% (93.2-98.1%) |
| **Projections** |  |  |  |  |  |
| AP/PA without lateral | 108 | 576 | 0.987 (0.979-0.993) | 93.5% (88.9-98.1%) | 95.0% (93.1-96.7%) |
| AP/PA with lateral | 20 | 281 | 0.990 (0.979-0.998) | 100.0% (100.0-100.0%) | 96.1% (94.0-98.2%) |

### Detection of pneumothorax and tension pneumothorax when ancillary findings present

The AI model accuracy was assessed in the presence or absence of ten ancillary findings. It mostly achieved AUC 0.95, sensitivity 80% and specificity 80% for the detection of both pneumothorax and tension pneumothorax in the presence or absence of each finding (Table 4). The exceptions included performing with AUC <0.95 and specificity <80% for the detection of pneumothorax in the presence of findings commonly associated with pneumothorax including rib fracture, pneumomediastinum, subcutaneous emphysema and intercostal drain; it also had AUC <0.95 for the detection of pneumothorax when there was evidence of thoracic surgery. In these situations, the model still performed with AUC >0.90.

**Table 4:**
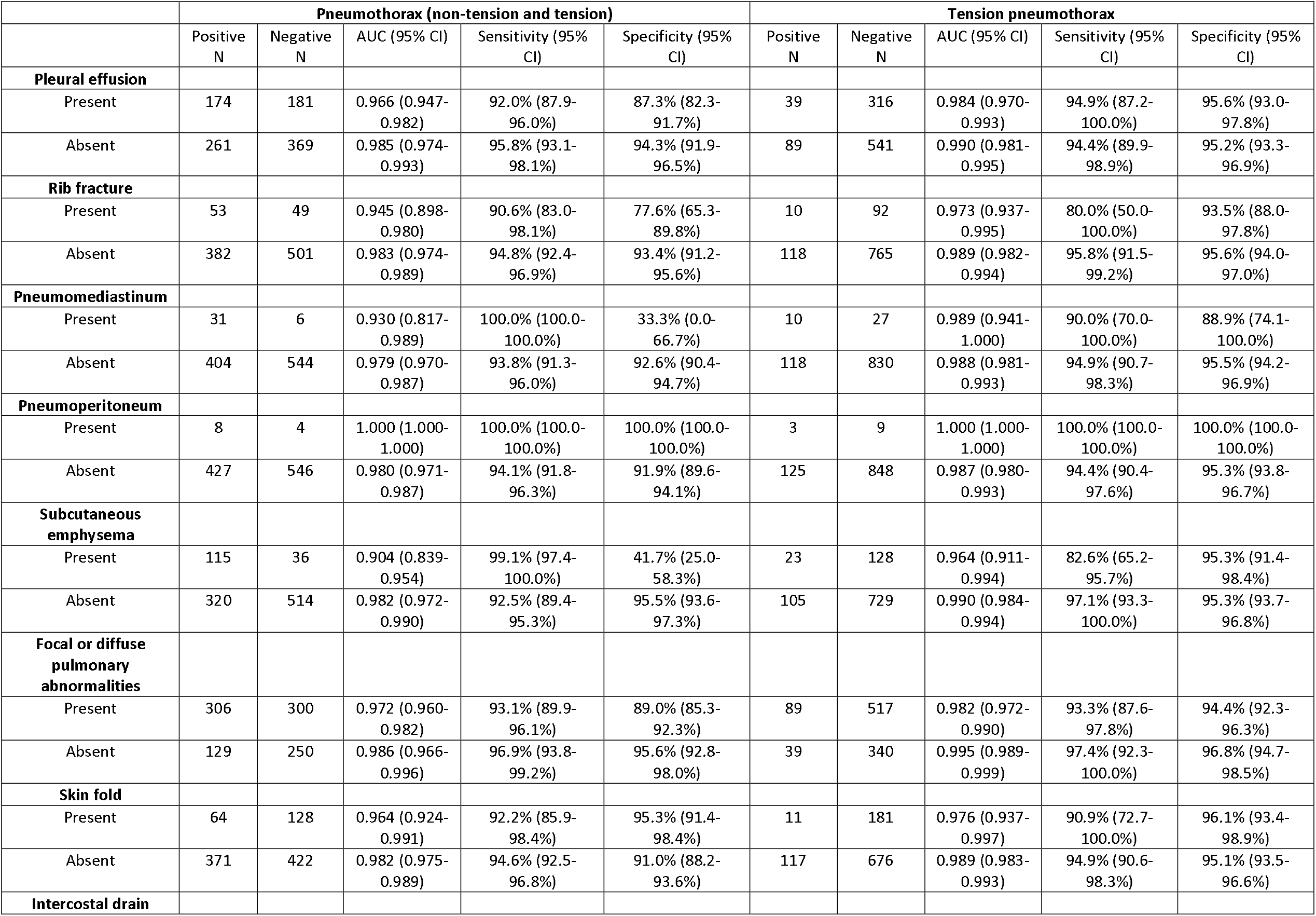
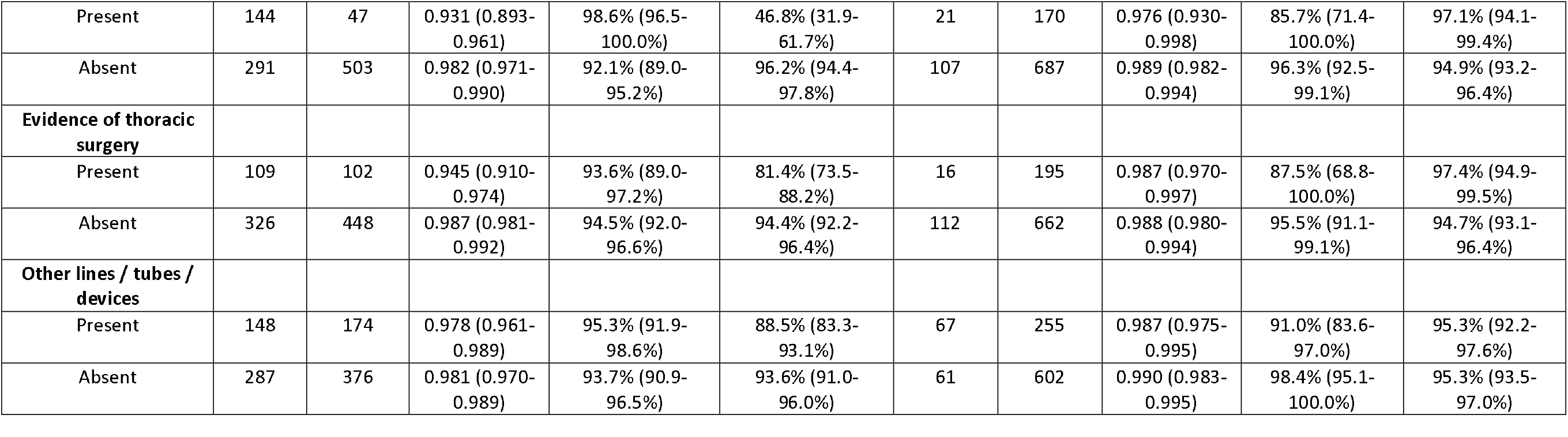
Performance for detecting pneumothorax (non-tension and tension) and tension pneumothorax in the presence of different ancillary findings.

## Discussion

This retrospective study assessed the performance of an AI model at detecting pneumothorax and tension pneumothorax on CXR. The model overall achieved AUC >0.95, sensitivity >80% and specificity >80%. These results are consistent with other FDA-cleared pneumothorax detection models.^10-14,16,17^ This model is, however, the first FDA-cleared model that also incorporates tension pneumothorax detection.

This study demonstrated consistent accuracy of the AI model in identifying pneumothorax and tension pneumothorax across demographic and technical subgroups including age, sex, manufacturer, patient positioning and CXR projection. This performance suggests that the model has generalizability for the diverse clinical scenarios that it may encounter moving forward. It also achieved sensitivity >80% for detecting pneumothorax when the pneumothorax size was <2cm or ≥2cm, and when the features of tension pneumothorax were present or absent. It unsurprisingly had a higher sensitivity for detecting a pneumothorax when it was larger (≥2cm) or had features of tension; it had sensitivity of 100% for the latter.

The AI model similarly demonstrated consistent model accuracy across most ancillary findings. There were, however, some ancillary findings that the model performed less well on. In particular, when the ancillary findings pneumomediastinum, subcutaneous emphysema and intercostal drain were present, the model had specificity of <50% for detection of pneumothorax. These three findings are commonly associated with pneumothorax and it is possible that the model uses them to identify pneumothorax; hence their presence could contribute to the model calling a case positive. It is also possible that their presence suggests a resolved pneumothorax that the model calls positive. A follow-on experiment could utilize the segmentation function of this model to understand where the model believes the pneumothorax is; this study utilized the subsequently FDA-cleared version that included the classification functionality and did not include the segmentation functionality. Similarly, the presence of the ancillary findings rib fracture and evidence of thoracic surgery each had AUC 0.945 for detection of pneumothorax; these results likely reflect the associations of these findings with pneumothorax albeit not as strong as the associations of pneumomediastinum, subcutaneous emphysema and intercostal drain.

The cohort selection included two strategies: the consecutive pneumothorax cases were identified by a manual review of the original radiology reports from consecutive CXR cases, while the tension pneumothorax cases were initially identified with natural language processing before the manual review. The first strategy ensured that cases reflected the distribution of real-world pneumothorax cases, including cases with a spectrum of pneumothorax conspicuity. The second strategy was used given the infrequency of tension pneumothorax amongst all CXR cases. One item that we noted was that there were fewer ground truth-positive cases compared to the number of original radiology report-positive cases. We hypothesized that this discrepancy occurred because of the lack of associated clinical history and the lack of other imaging such as chest computed tomography, which might have increased the detection rate in the original clinical environment. The ground truth radiologists in this current study only had access to the CXR.

A key limitation of this study is that it is a retrospective study outside of the clinical workflow. While it demonstrates the accuracy of the AI model in interpreting imaging across many demographic and technical subgroups, it does not do so within the broader clinical environment. Further evaluation will be required to know how the model impacts the clinical workflow including case prioritization and patient outcomes. Further evaluation will also be required as the model encounters clinical scenarios beyond the current study including from new radiographic equipment manufacturers or models.

## Conclusion

This study assessed an AI model that accurately detected pneumothorax and tension pneumothorax. Its use in the clinical environment may lead to earlier identification and improved care for patients with pneumothorax.

## Data Availability

The AI model is part of the FDA cleared Annalise Enterprise CXR Triage Pneumothorax device, which is commercially available in the US. The test dataset generated for this study contains protected patient information. Some data may be available for research purposes from the corresponding author upon reasonable request.

## Acknowledgements

The authors thank the broader Mass General Brigham Data Science Office and Annalise teams for their assistance with this project.

