## Supplementary figures and images for "Evaluation of an artificial intelligence model for detection of pneumothorax and tension pneumothorax on chest radiograph"

### Supplementary Figure 1

**Supplementary Figure 1: Case selection flow chart**

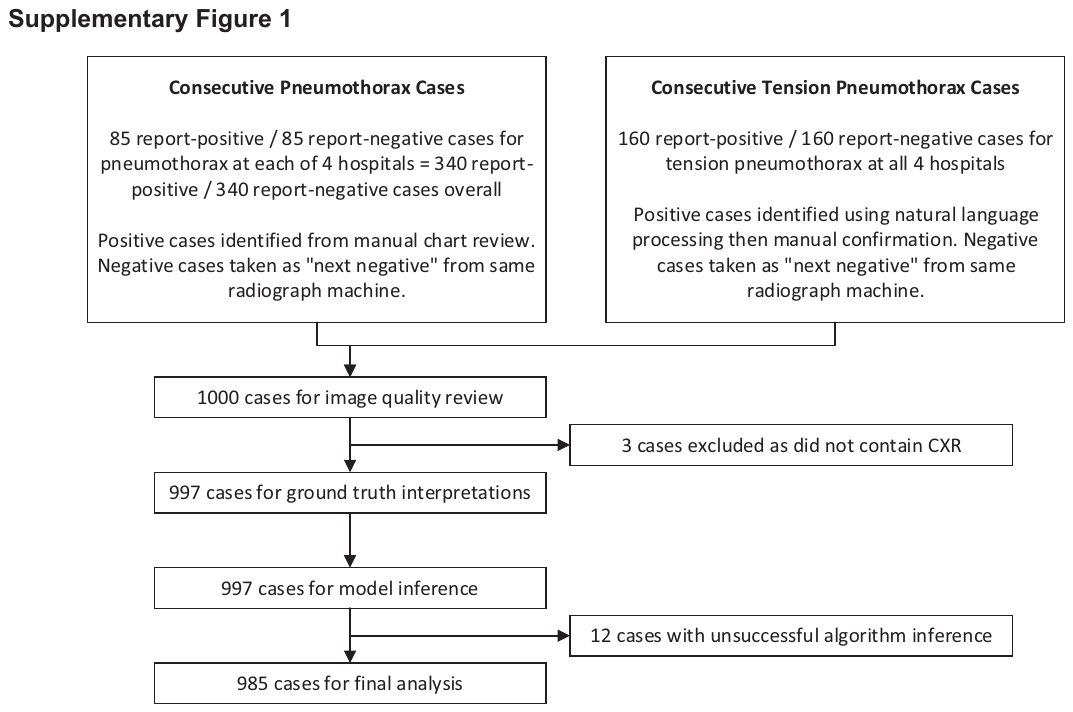
